# Reduced magnitude and durability of humoral immune responses by COVID-19 mRNA vaccines among older adults

**DOI:** 10.1101/2021.09.06.21263149

**Authors:** Mark A. Brockman, Francis Mwimanzi, Hope R. Lapointe, Yurou Sang, Olga Agafitei, Peter Cheung, Siobhan Ennis, Kurtis Ng, Simran Basra, Li Yi Lim, Fatima Yaseen, Landon Young, Gisele Umviligihozo, F. Harrison Omondi, Rebecca Kalikawe, Laura Burns, Chanson J. Brumme, Victor Leung, Julio S.G. Montaner, Daniel Holmes, Mari DeMarco, Janet Simons, Ralph Pantophlet, Masahiro Niikura, Marc G. Romney, Zabrina L. Brumme

## Abstract

**Background:** mRNA vaccines reduce COVID-19 incidence and severity, but the durability of vaccine-induced immune responses, particularly among the elderly, remains incompletely characterized.

**Methods:** Anti-spike RBD antibody titers, ACE2 competition and virus neutralizing activities were longitudinally assessed in 151 healthcare workers and older adults (overall aged 24-98 years) up to three months after vaccination.

**Results:** Older adults exhibited lower antibody responses after one and two vaccine doses for all measures. In multivariable analyses correcting for sociodemographic, chronic health and vaccine-related variables, age remained independently associated with all response outcomes. The number of chronic health conditions was additionally associated with lower binding antibody responses after two doses, and male sex with lower ACE2 competition activity after one dose. Responses waned universally at three months after the second dose, but binding antibodies, ACE2 competition and neutralizing activities remained significantly lower with age. Older adults also displayed reduced ability to block ACE2 binding by the Delta variant.

**Conclusions:** The humoral immune response to COVID-19 mRNA vaccines is significantly weaker with age, and universally wanes over time. This will likely reduce antibody-mediated protection against SARS-CoV-2 and the Delta variant as the pandemic progresses. Older adults may benefit from additional immunizations as a priority.

## INTRODUCTION

Older age is the greatest risk factor for lethal coronavirus disease 2019 (COVID-19) following infection with severe acute respiratory syndrome coronavirus 2 (SARS-CoV-2) ^1-3^. While COVID-19 vaccines offer hope to end the pandemic ^4-7^, “real world” assessments have revealed weaker vaccine-induced immune responses in certain groups including the elderly ^8-13^, though few studies have adjusted for potential confounders, including comorbidities, that can accumulate with age. Vaccine response durability also remains incompletely characterized, as immunogenicity assessments are occurring concomitantly with national vaccine rollouts.

Two mRNA vaccines, BNT162b2 (Pfizer/BioNTech) and mRNA-1273 (Moderna), have been administered widely in North America and Europe. While both provided >94% protection against moderate or severe COVID-19 in clinical trials after two doses ^6,7^, and while population-level reductions in COVID-19 were clearly observed following initial vaccine rollouts, ongoing outbreaks in long-term care facilities underscore the continuing vulnerability of older adults to SARS-CoV-2 infection, even after vaccination ^14^. Longitudinal assessments of the magnitude and durability of vaccine-induced immune responses can inform public health decision-making for older adults as the pandemic progresses. However, while age and age-associated comorbidities, including chronic health conditions that can result in immune dysregulation, have been linked to poor vaccine immune responses ^15-17^, few studies have explored these variables in the context of COVID-19 immunization.

We investigated the magnitude of SARS-CoV-2-specific humoral immune responses after one and two mRNA vaccine doses in 151 participants aged 24-98 years. In a subset of participants with specimens available at time of writing, we further examined response durability three months following the second vaccine dose, including against the widely circulating Delta variant (B.1.617.2). Our results demonstrate weaker humoral responses to COVID-19 mRNA vaccines in older versus younger adults, signified by lower magnitude and durability of spike-specific binding and neutralizing antibody activities even after correction for potential confounders, and potentially reduced neutralization of the Delta variant.

## METHODS

### Study design

We conducted a prospective longitudinal cohort study in British Columbia, Canada, to examine SARS-CoV-2 specific humoral immune responses following COVID-19 mRNA vaccination. The cohort, totaling 151 individuals, comprised 89 healthcare workers and 62 older adults (including 23 residents of long-term care and assisted living facilities, and 39 seniors living independently).

### Ethics approval

Written informed consent was obtained from all participants or their authorized substitute decision makers. This study was approved by the University of British Columbia/Providence Health Care and Simon Fraser University Research Ethics Boards (protocol H20-03906).

### Participants and sampling

Participants were recruited at facilities operated by Providence Health Care (Vancouver) and the community. Serum and plasma were collected prior to vaccination, one month after the first dose, and at one and three months after the second dose. Specimens were processed same-day and frozen until analysis. COVID-19 convalescent individuals were identified at study entry by the presence of serum antibodies recognizing SARS-CoV-2 nucleoprotein (N).

### Data sources and immunity measures

Sociodemographic data (age, sex, ethnicity), chronic health conditions and COVID-19 vaccination information were collected by self-report and confirmed through medical records where available. Chronic health conditions were defined as hypertension, diabetes, chronic diseases of lung, liver, kidney, heart or blood, cancer and immunosuppressive conditions/drugs to create a numerical variable (0-9 conditions) per participant. Vaccine-induced antibody responses against SARS-CoV-2 were assessed three ways: (1) Commercial and in-house assays to detect binding antibodies targeting the spike receptor binding domain (RBD); (2) Angiotensin-converting enzyme 2 (ACE2) competition assays to detect receptor-blocking antibodies; and (3) Neutralization assays to detect antibodies that prevent virus infection of target cells.

### Binding antibody assays

We examined total binding antibodies against the SARS-CoV-2 N and RBD in serum using the Elecsys Anti-SARS-CoV-2 assay (which detects anti-N antibodies generated following infection) and Anti-SARS-CoV-2 S assay (which quantifies total antibodies against RBD generated following infection or vaccination), respectively, on a Cobas e601 module analyzer (Roche Diagnostics). Both assays incorporate sandwich ELISA and electro-chemiluminescence detection and report results in arbitrary units/mL, calibrated against an external standard. Sera were tested undiluted, with samples above the upper limit of quantification re-tested at 1:100 dilution. We also quantified the IgG sub-component of the plasma antibody response to RBD using bead-based ELISA on a Luminex 200 instrument. Here, recombinant His-tagged RBD (Wuhan or Delta/B.1.617.2 strain; R&D Systems) was coupled to carboxylated xMAP beads (Bio-Rad). Plasma was tested in duplicate following 1:200 or 1:3200 dilution. Bound IgG was detected using PE-conjugated anti-human IgG secondary antibody (BioLegend) with results reported as arbitrary median fluorescence intensities (MFI).

### ACE2 competition assay

We assessed the ability of plasma antibodies to block the interaction between RBD and the ACE2 receptor using competition ELISA on a Luminex 200 instrument. Here, plasma samples were diluted 1:100 in buffer containing a non-saturating concentration of soluble recombinant biotinylated ACE2 (50 nM; Abcam) prior to incubation with RBD-coupled xMAP beads. Bound ACE2 was detected using streptavidin-PE (Bio-Rad). ACE2 displacement was calculated as 100 - [MFI in the presence of plasma /MFI in the absence of plasma] and reported as a percentage.

### Virus neutralization assays

Neutralizing activity in plasma was examined using a live SARS-CoV-2 infectivity assay in a Containment Level 3 facility. Assays were performed using isolate USA-WA1/2020 (BEI Resources) and VeroE6-TMPRSS2 (JCRB-1819) target cells. Viral stock was adjusted to 50 TCID_50_/200 µl in DMEM in the presence of serial 2-fold dilutions of plasma (initial dilution, 1:20), incubated at 4°C for 1 hour and then added to target cells in 96-well plates in triplicate. The appearance of viral cytopathic effect (CPE) was recorded 3 days post-infection. Neutralizing activity is reported as “present” if CPE was prevented in at least one of three replicate wells at a 1/20 or higher plasma dilution (binary variable); or as the reciprocal plasma dilution necessary to prevent CPE in all triplicate wells (continuous variable).

### Statistical analysis

Comparisons of binary variables between groups were performed using Fisher’s exact test. Comparisons of continuous variables between groups were performed using the Mann-Whitney U-test (for unpaired data) or Wilcoxon test (for paired data). Ordinary least squares regression was used to examine relationships between continuous variables. Multiple linear regression was employed to investigate the relationship between age (per year increment), sex (female as reference group), Ethnicity (non-white as reference group), number of chronic health conditions (per number increment), vaccine type (Pfizer as reference group) and sampling date following vaccine dose (per day increment) on immunogenicity outcomes. All tests were two-tailed, with p=0.05 considered statistically significant. Analyses were conducted using Prism v9.2.0 (GraphPad).

## RESULTS

### Lower RBD binding antibodies associated with older age and number of chronic health conditions

Characteristics of the 151 participants, which included 89 healthcare workers (HCW) and 62 older adults (Seniors+LTC) are shown in **Table 1**. All participants received two doses of an mRNA vaccine between December 2020-July 2021. Due to limited initial vaccine supply in Canada, the interval between first and second doses was extended to a maximum of 112 days in British Columbia beginning on March 1, 2021, so participants received their second dose a median of 91 days after the first (interquartile range [IQR] 70-99 days). Samples were collected before vaccination to assess prior exposure to SARS-CoV-2 (n=142); at one month following the first (n=141) and second (n=147) doses to quantify response magnitude; and at three months following the second dose (n=30) to examine response durability.

**Table 1.** Study Participants (n=151)

|  | **Health Care Workers (n=89)** | **Seniors + LTC (n=62)** | **p-value** |
| --- | --- | --- | --- |
| **Age in years, median [IQR]** | 41 [35-50] | 79 [73-86] | **<0.0001** |
| **Female Sex, n (%)** | 65 (73%) | 43 (69%) | 0.71 |
| **White/Caucasian Ethnicity, n (%)** | 40 (45%) | 47 (76%) | **0.0002** |
| **COVID-19 convalescent (anti-N Ab+), n (%)** | 8 (9%) | 6 (10%) | >0.99 |
| **Chronic health or immunosuppressive conditions, median [IQR]** | 0 [0-0] | 1 [0-2] | **<0.0001** |
| **Pfizer mRNA Vaccine, n (%)** | 88 (99%) | 54 (87%) | **0.0015** |
| **Time between doses in days, median [IQR]** | 97 [91-103] | 78 [45-86] | **<0.0001** |
| **Specimens collected one month after first dose, n (%)** | 87 (98%) | 54 (87%) | n.a. |
| **Day of specimen collection one month after first dose, median [IQR]** | 29 [27-31] | 30 [28-32] | **0.0069** |
| **Specimens collected one month after second dose, n (%)** | 89 (100%) | 58 (87%) | n.a. |
| **Day of specimen collection one month after second dose, median [IQR]** | 30 [29-32] | 30 [29-32] | 0.96 |
| **Specimens collected three months after second dose, n (%)** | 13 (15%) | 17 (27%) | n.a. |
| **Day of specimen collection three months after second dose, median [IQR]** | 91 [90-95] | 90 [90-91] | 0.19 |

IQR, interquartile range; n.a., not applicable

HCW and older adults (Seniors+LTC) were a median of 41 and 79 years old respectively, and predominantly female. At study entry, 14 participants (9.3%; eight HCW and six older adults) were identified as COVID-19 convalescent based on the presence of anti-SARS-CoV-2 N antibodies. Nine participants (6%; one HCW and eight older adults) received the Moderna vaccine for their first dose, while 142 (94%) received Pfizer. In addition to age, the two groups differed significantly in terms of ethnicity (p=0.0002), number of chronic health conditions (p<0.0001), vaccine received (p=0.0015), time between doses (p<0.0001) and specimen collection date after the first dose (p=0.0069).

We quantified interim and peak anti-RBD IgG binding antibodies in plasma one month after the first and second vaccine doses using a bead-based Luminex ELISA assay. After one dose, median anti-RBD IgG titers were 4.2-fold lower in Seniors+LTC compared to HCW (p<0.0001) (**Figure 1A**). In contrast, the 14 COVID-19 convalescent participants mounted ∼190- and ∼840-fold higher IgG responses after one dose compared to COVID-19 naïve HCW and Seniors+LTC, respectively (both p<0.0001), consistent with prior studies demonstrating robust reactivity to one dose in previously infected individuals ^18^. After two doses, median anti-RBD IgG responses increased by a median of >2 log in both naïve groups (**Figure 1C; Supplemental Figure 1A**), with some indication that the extent of boosting was marginally higher in older individuals (**Supplemental Figure 1B**), but no further increase was observed in convalescent participants. Indeed, after two doses the median IgG values in HCW reached equivalence with the convalescent group, while values in Seniors+LTC remained ∼3-fold lower (both comparisons p<0.0001), with two doubly-vaccinated older adults continuing to exhibit very poor responses (**Figure 1C**). No difference in binding antibody responses was seen between Seniors and LTC residents after one or two vaccine doses when these groups were analyzed separately (both p>0.3; not shown), supporting their analysis as a single group of older adults.

**Figure 1:**
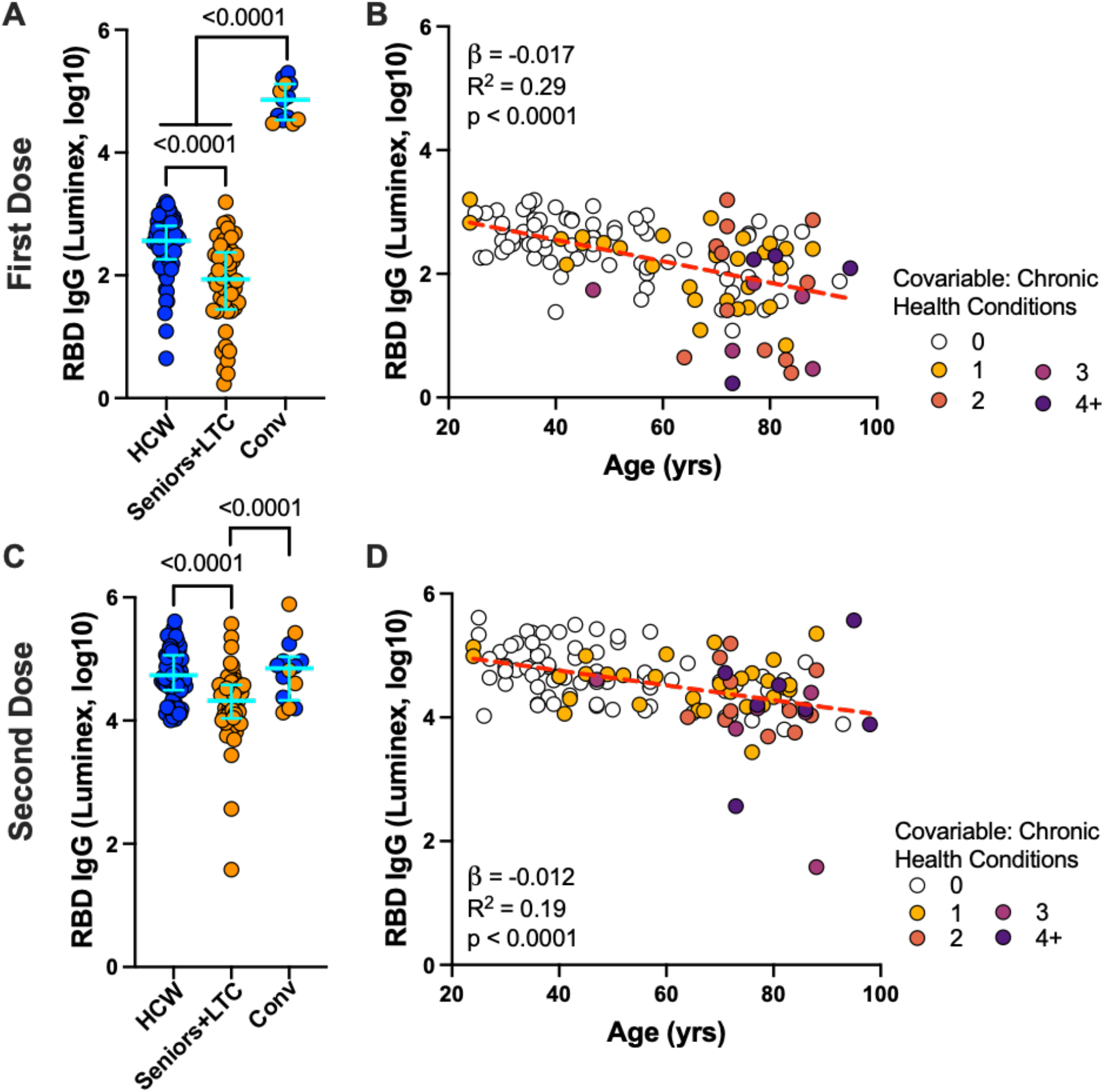
Binding antibody responses to spike RBD following one and two doses of a COVID-19 mRNA vaccine are lower in older adults. *Panel A*: Binding IgG responses to the SARS-CoV-2 spike RBD in plasma, measured using a Luminex ELISA assay, following one dose of a COVID-19 mRNA vaccine in Seniors+LTC (comprising individuals living in long-term care or assisted living facilities and seniors living independently; orange circles) and healthcare workers (HCW; blue circles), who were COVID-19 naive at study entry. The third group, convalescents (“Conv”) denotes participants with anti-N antibodies at study entry, colored as above. Bars represent median and IQR. P-values were computed using the Mann-Whitney U-test. *Panel B*: Same data as the HCW and Seniors+LTC groups shown in panel A, but plotted by age, and where participants are colored based on their number of chronic health conditions, which remained significant in multivariable analyses (see Table 2). Statistics computed using ordinary least-squares regression, also shown as dotted line. *Panels C, D:* Same as A and B, but for responses following two doses of mRNA vaccine.

Among COVID-19 naïve individuals, we estimate using univariable linear regression that every 10 years of older age was associated, on average, with 0.17 and 0.12 log_10_ lower IgG responses, after one and two vaccine doses, respectively (**Figures 1B,D**). Multivariable analyses in COVID-19 naïve individuals adjusting for sex (as studies have reported enhanced immune function in females following pathogen infection and vaccination ^19^), ethnicity, number of chronic health conditions, vaccine brand, and day of specimen collection following immunization confirmed that age remained significantly associated with IgG responses after one and two vaccine doses (p=0.0001 and p=0.0005, respectively), and that the number of chronic health conditions also contributed negatively to these outcomes (p=0.005 and p=0.05, respectively) (**Table 2**). Note that the multivariable model for the response after two vaccine doses did not include the dosing interval as this variable was collinear with our age-defined participant groupings (the median dosing interval was lower in Senior+LTC compared to HCW because most LTC residents received two doses before BC extended the dose interval to 112 days). The lack of relationship between dosing interval and IgG response *within* HCW and Seniors+LTC groups (whose dose intervals ranged from 30-122 and 42-102 days, respectively, both p>0.6, not shown) provided additional support for excluding this variable from the model.

**Table 2.** Multivariable Analyses

| Immunogenicity outcome | Variable | Time point |  |  |  |
| --- | --- | --- | --- | --- | --- |
|  |  | 1 month after 1st dose |  | 1 month after 2nd dose |  |
| | | $\beta$ estimate (95% CI) | p | $\beta$ estimate (95% CI) | p |
| Log10 RBD IgG | Age | -0.012 (-0.018 to -0.0061) | **0.0001** | -0.0010 (-0.015 to -0.0045) | **0.0005** |
|  | Male Sex | -0.12 (-0.33 to 0.086) | 0.2 | -0.10 (-0.29 to 0.090) | 0.3 |
|  | White ethnicity | -0.070 (-0.28 to 0.14) | 0.5 | 0.034 (-0.15 to 0.22) | 0.7 |
|  | # Chronic health conditions | -0.15 (-0.25 to -0.045) | **0.005** | -0.084 (-0.17 to -0.00042) | **0.05** |
|  | Moderna vaccine (vs. Pfizer) | -0.22 (-0.61 to 0.18) | 0.3 | 0.24 (-0.11 to 0.59) | 0.2 |
|  | Days after vaccine dose | 0.00085 (-0.037 to 0.039) | 1 | -0.0071 (-0.043 to 0.029) | 0.7 |
| % ACE2 displacement | Age | -0.15 (-0.30 to -0.0063) | **0.04** | -0.38 (-0.55 to -0.21) | **<0.0001** |
|  | Male Sex | -6.51 (-11.7 to -1.33) | **0.01** | -4.60 (-10.36 to 1.15) | 0.1 |
|  | White ethnicity | -1.97 (-7.073 to 3.13) | 0.4 | 0.90 (-4.84 to 6.63) | 0.8 |
|  | # Chronic health conditions | -0.52 (-3.059 to 2.011) | 0.7 | -2.36 (-4.76 to 0.21) | 0.07 |
|  | Moderna vaccine (vs. Pfizer) | 2.45 (-7.34 to 12.24) | 0.6 | 5.79 (-4.85 to 16.44) | 0.3 |
|  | Days after vaccine dose | -0.48 (-1.43 to 0.46) | 0.3 | -0.27 (-1.36 to 0.82) | 0.6 |
| Viral Neutralization<br>(log2 plasma dilution factor) | Age | -0.0049 (-0.0087 to -0.0010) | **0.01** | -0.036 (-0.060 to -0.011) | **0.006** |
|  | Male Sex | -0.083 (-0.22 to 0.050) | 0.2 | -0.97 (-2.04 to 0.087) | 0.07 |
|  | White ethnicity | -0.075 (-0.20 to 0.056) | 0.3 | -0.15 (-1.38 to 1.078) | 0.8 |
|  | # Chronic health conditions | 0.033 (-0.027 to 0.094) | 0.3 | 0.051 (-0.25 to 0.36) | 0.7 |
|  | Moderna vaccine (vs. Pfizer) | 0.11 (-0.19 to 0.42) | 0.5 | -1.06 (-2.91 to 0.80) | 0.3 |
|  | Days after vaccine dose | -0.0030 (-0.027 to 0.021) | 0.8 | 0.022 (-0.24 to 0.29) | 0.9 |

CI, confidence interval

Highly comparable results were obtained from sera using the commercial Roche Elecsys Anti-SARS-CoV-2 S assay, which assesses total anti-RBD antibodies (**Supplemental Figure 2 and Supplemental Table 1**), confirming that the binding antibody response elicited by COVID-19 mRNA vaccines is significantly lower in older adults, even after two immunizations.

### Lower ability to block ACE2 receptor binding is associated with older age and male sex

We next assessed the ability of plasma to block the RBD-ACE2 interaction using Luminex ELISA, a higher throughput approach to estimate potential viral neutralization activity. After one vaccine dose, HCW and Seniors+LTC exhibited median 45% and 38% ACE2 displacement activities, respectively, indicating lower function among older adults (p=0.0026) (**Figure 2A**). In contrast, convalescent participants exhibited a median 92% ACE2 displacement activity after one dose (p<0.0001 compared to both naive groups). Following two vaccine doses, HCW exhibited a median 91% ACE2 displacement activity compared to a median of 70% in Seniors+LTC (p<0.0001) (**Figure 2C**). Of note, while the second dose boosted *binding antibodies* to similar (or higher) levels in Seniors+LTC compared to HCW (**Supplemental Figure 1A, B**), it had significantly less of an impact on the ability of these antibodies to displace ACE2 in Seniors+LTC compared to HCW (p=0.0003) (**Supplemental Figure 1C**). This suggests that, though these mRNA vaccines clearly stimulate antibody production, antibody functions such as affinity maturation may be diminished with age.

**Figure 2:**
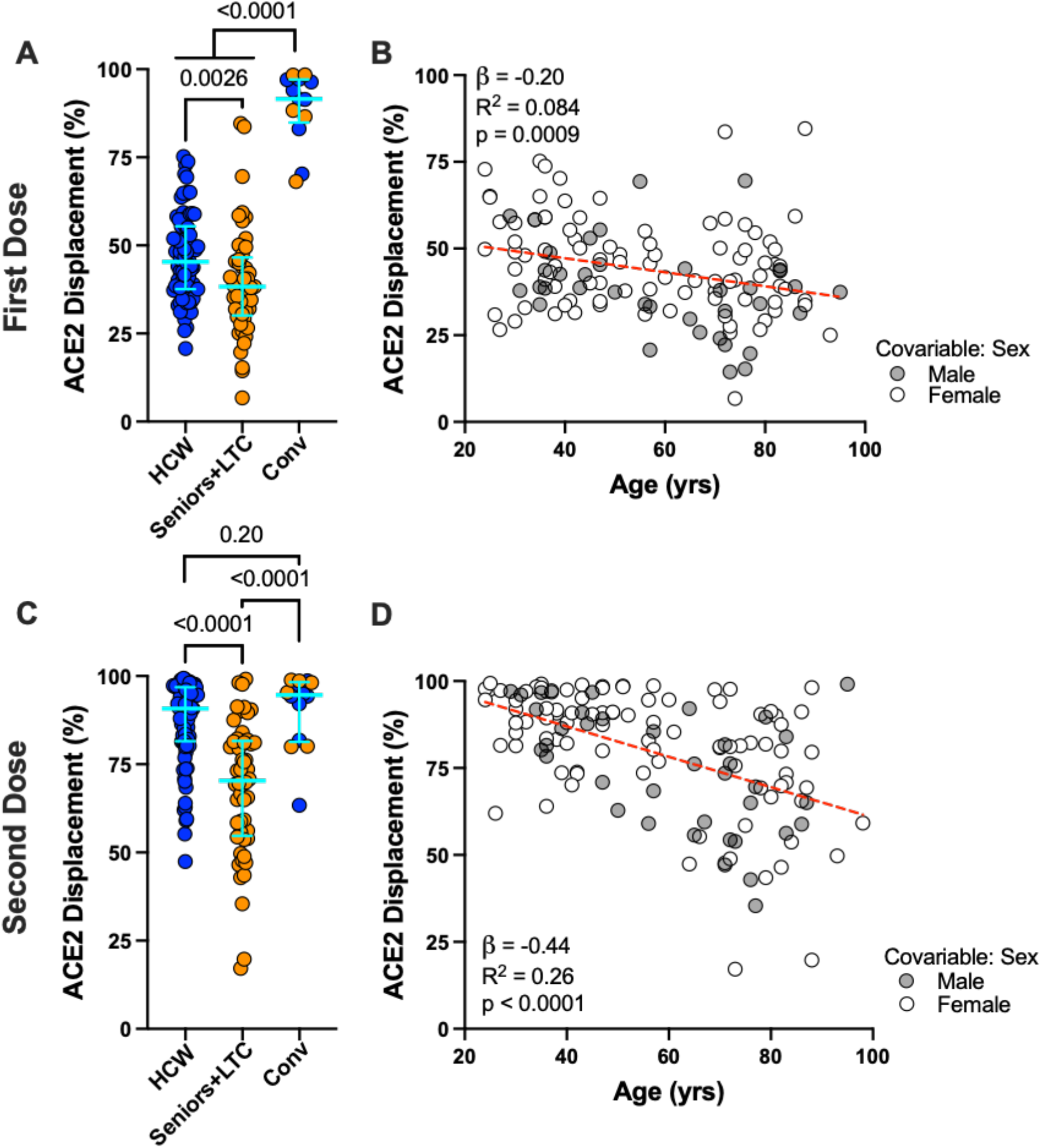
Ability of vaccine-induced antibodies to block ACE2-receptor binding is weaker in older adults. *Panel A*: Ability of vaccine-induced plasma antibodies to displace soluble ACE2-receptor from spike RBD, measured using a Luminex ELISA assay, following one dose of vaccine in COVID-19 naive Seniors+LTC (orange circles), HCW (blue circles), and COVID-19 convalescent participants (“Conv”; colored as above). Bars represent median and IQR. P-values computed using the Mann-Whitney U-test. *Panel B*: Same data as the HCW and Seniors+LTC groups shown in panel A, but plotted by age, and colored by sex, which remained significant in multivariable analyses (see Table 2). Statistics computed using ordinary least-squares regression, also shown as dotted line. *Panels C, D:* Same as A and B, but for responses following two doses of mRNA vaccine.

Among COVID-19 naïve individuals, we estimate using univariable linear regression that every 10 years of older age was associated, on average, with 2.0% and 4.4% lower ACE2 displacement activity after one and two vaccine doses respectively (**Figures 2B,D**). Multivariable regression in COVID-19 naïve individuals confirmed that age remained associated with ACE2 displacement activity after one (p=0.04) and two (p<0.0001) doses (**Table 2**). Female sex was independently associated with 6.5% higher ACE2 displacement activity after one dose (p=0.01), which is notable since women have been reported to display higher neutralizing antibody responses following infection and vaccination ^19^. Chronic health conditions also played a potential negative role on ACE2 displacement activity after two doses (p=0.07).

### Weaker virus neutralizing activity is associated with age and chronic health conditions

We next performed SARS-CoV-2 neutralization assays to quantify the ability of plasma to block virus infection of target cells, which may involve spike epitopes outside of the RBD. Following one vaccine dose, 15 of 72 (21%) COVID-19 naïve HCW displayed modest evidence of neutralizing activity (at 1:20 dilution), compared to only one of 32 (3%) Seniors+LTC (p=0.02) (**Figure 3A**), a relationship that also held when age was analyzed as a continuous variable (**Figure 3B**). In contrast, all 13 convalescent participants efficiently neutralized live SARS-CoV-2 following one vaccine dose (median reciprocal titer of 240) (**Figure 3A**), despite showing no neutralization activity at 1:20 plasma dilution prior to vaccination (not shown). Following the second vaccine dose, all 15 tested HCW displayed neutralizing activity at 1:20 dilution compared to only 11 of 17 (65%) older adults (p=0.009; **Figure 3C**). Consistent with ACE2 displacement results, the second vaccine dose boosted virus neutralization titers significantly better in HCW compared to Seniors+LTC (**Supplemental Figure 1E,F**). Indeed, ACE2 displacement activity correlated with virus neutralization activity (Spearman ρ=0.82; p<0.0001), suggesting that the former is a reasonable surrogate for the latter. In univariate analyses of COVID-19 naïve individuals, we estimate that every 10 years of older age was associated with an average 0.4 log2 lower neutralization after two doses (**Figure 3D**). In multivariable analyses in COVID-19 naïve individuals, age remained the only significant contributor to virus neutralization activity after one and two vaccine doses (p=0.01 and p=0.006, respectively) (**Table 2**).

**Figure 3:**
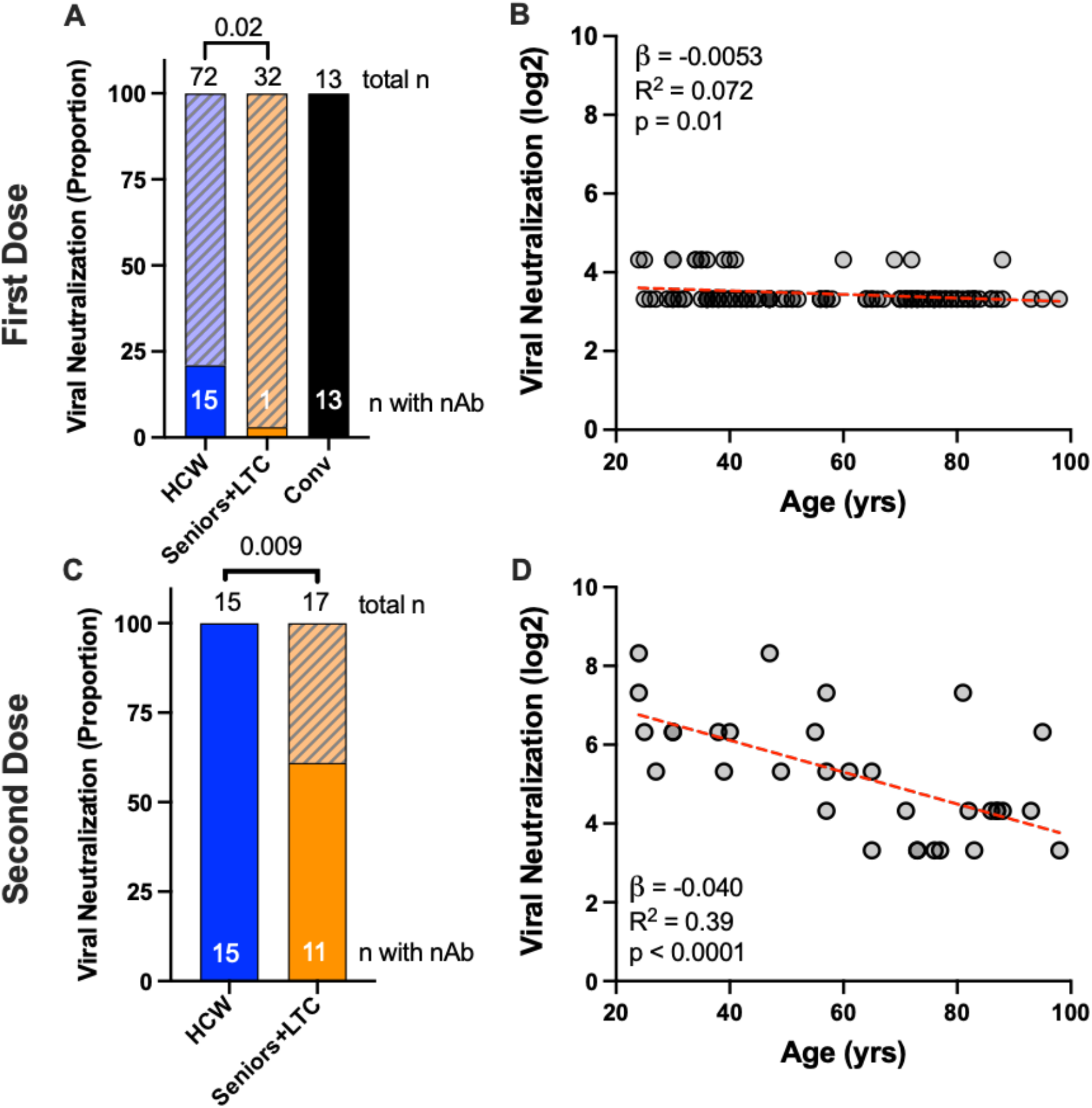
Viral neutralization activity of vaccine-induced antibodies is reduced in older adults. *Panel A*: The proportion of participants in each group who displayed any neutralizing activity against live SARS-CoV-2 (USA-WA1/2020 strain) are shown as the solid part of each histogram, where COVID-19 naive HCW and Seniors+LTC are shown in blue and orange respectively, and convalescent participants (Conv) are shown in black. Total Ns in each group are shown above each bar, with the N of participants displaying neutralizing activity shown within the bar. P-values computed using Fisher’s exact test. *Panel B*: Same data as the HCW and Seniors+LTC groups shown in panel A, but plotted by age, and where neutralization is reported as the reciprocal log2 dilution value. Samples that displayed neutralization in fewer than three wells at a 1/20 dilution were coded as having a reciprocal dilution factor of 10 (3.32 log 2). Statistics computed using ordinary least-squares regression. *Panels C, D*: Same as A and B, but for neutralization responses after two doses of mRNA vaccine in a subset of participants.

### Vaccine-induced antibody responses decline over time across all ages

At time of writing, plasma from 30 COVID-19 naive participants (13 HCW and 17 LTC residents, median 38 and 82 years old respectively) were available at three months following two vaccine doses, allowing us to assess vaccine response durability. In essentially all participants, antibody responses declined markedly between one and three months following the second dose: median IgG binding antibodies declined 7-fold and 4.5-fold in HCW and LTC, respectively (both p≤0.0001; **Figure 4A**) while median ACE2 displacement activity declined by 39% in HCW (p=0.0002) and by 30% in LTC (p=0.003) (**Figure 4B**). Median virus neutralization activity declined 4-fold in HCW (p=0.0005) and 2-fold in LTC, though the latter did not reach statistical significance (p=0.06) (**Figure 4C**). Despite these near-universal temporal reductions, group-specific medians remained substantially higher in HCW compared to LTC participants in all assays, with median residual activities in HCW at three months after the second dose approximating the peak response in LTC one month after this dose.

**Figure 4:**
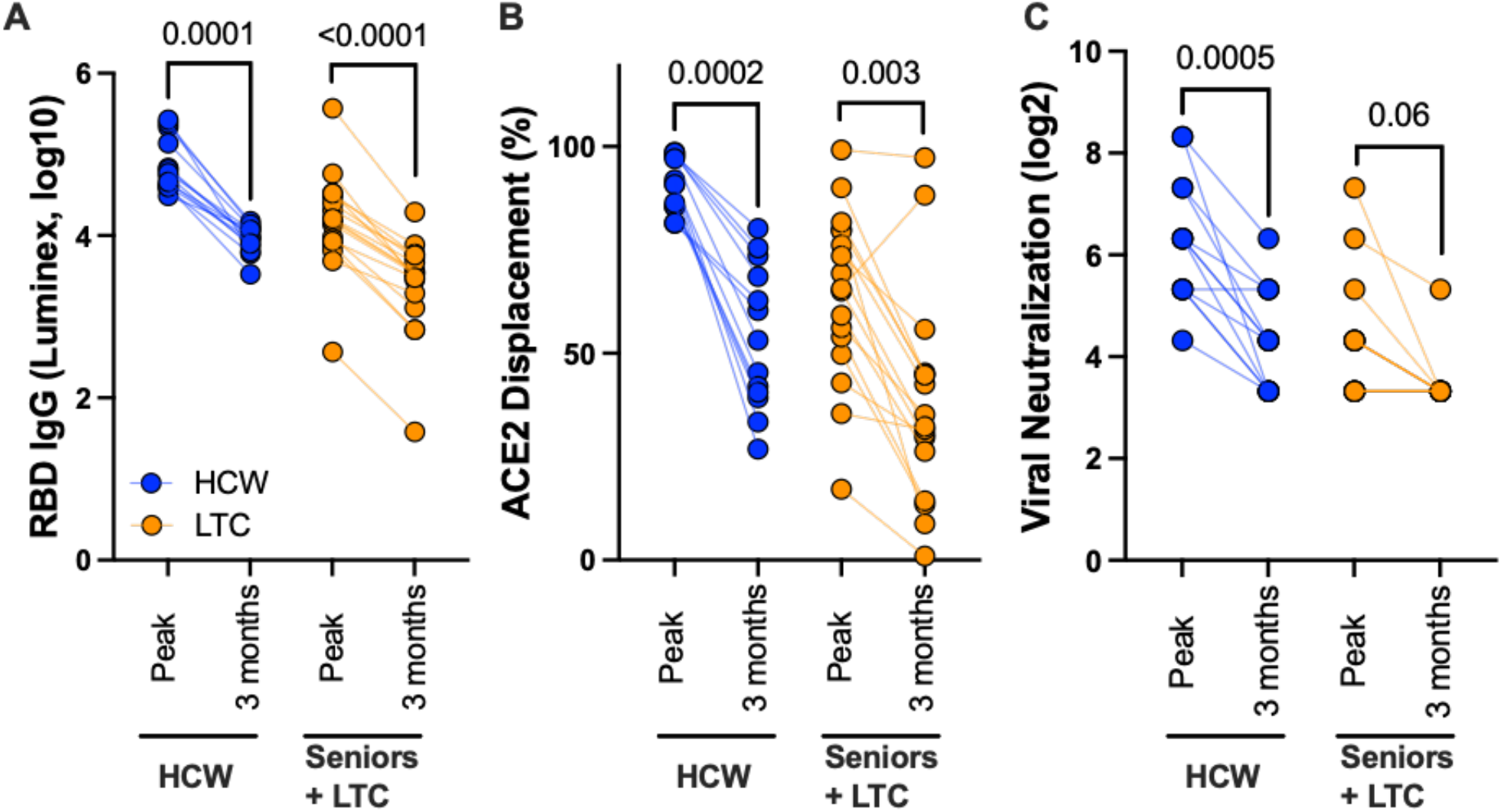
Waning of antibody responses after two doses of mRNA vaccine. *Panel A:* Binding IgG responses to spike RBD, measured by Luminex ELISA, one month following the second vaccine dose (peak) and three months after this dose (3 months) in a subset of HCW (blue circles) and individuals living in long-term care or assisted living facilities (LTC; orange) who were COVID-19 naive at study entry. P-values computed using the Wilcoxon paired test. *Panel B*: ACE2 competition assay results, measured by Luminex ELISA, in the same individuals. *Panel C*: Virus neutralization assay results, displayed as the reciprocal log 2 plasma dilution, in the same individuals. Note that some values are superimposed.

### Impaired ability to block ACE2 binding by Delta variant among older adults

Given recent concerns that certain SARS-CoV-2 variants may be more transmissible or evade aspects of host immunity ^20,21^, we examined binding antibodies and ACE2 competition activity against the widespread B.1.617.2 (Delta) variant at one and three months following the second vaccine dose in the same subset. Overall, anti-RBD binding antibody levels were equivalent between the original Wuhan strain and the Delta variant within each group and timepoint tested (all p>0.3; **Figure 5A**). Nevertheless, responses in LTC participants were lower than those in HCW at all timepoints, and responses in both groups waned considerably between one and three months after the second dose (**Figure 5A**). In contrast, plasma from both groups was impaired in its ability to block ACE2 receptor engagement by the Delta RBD compared to the Wuhan RBD: in HCW, median ACE2 displacement values were ∼7% lower against Delta at both one and three months after vaccination (both p≤0.0002), while in LTC, median ACE2 displacement values were ∼15% lower against Delta at both time points (both p≤0.0001) (**Figure 5B**). These results indicate that, while two vaccine doses can elicit binding antibodies that cross-recognize Delta RBD, these responses may be less able to prevent infection by this variant. This observation is consistent with a recent report showing reduced ability of plasma from convalescent and vaccinated individuals to neutralize this strain ^22^. Considering the lower peak immune response in older adults, combined with the waning of these responses over time, our data suggest that older adults will remain more susceptible to infection by the Delta variant even after two vaccine doses.

**Figure 5:**
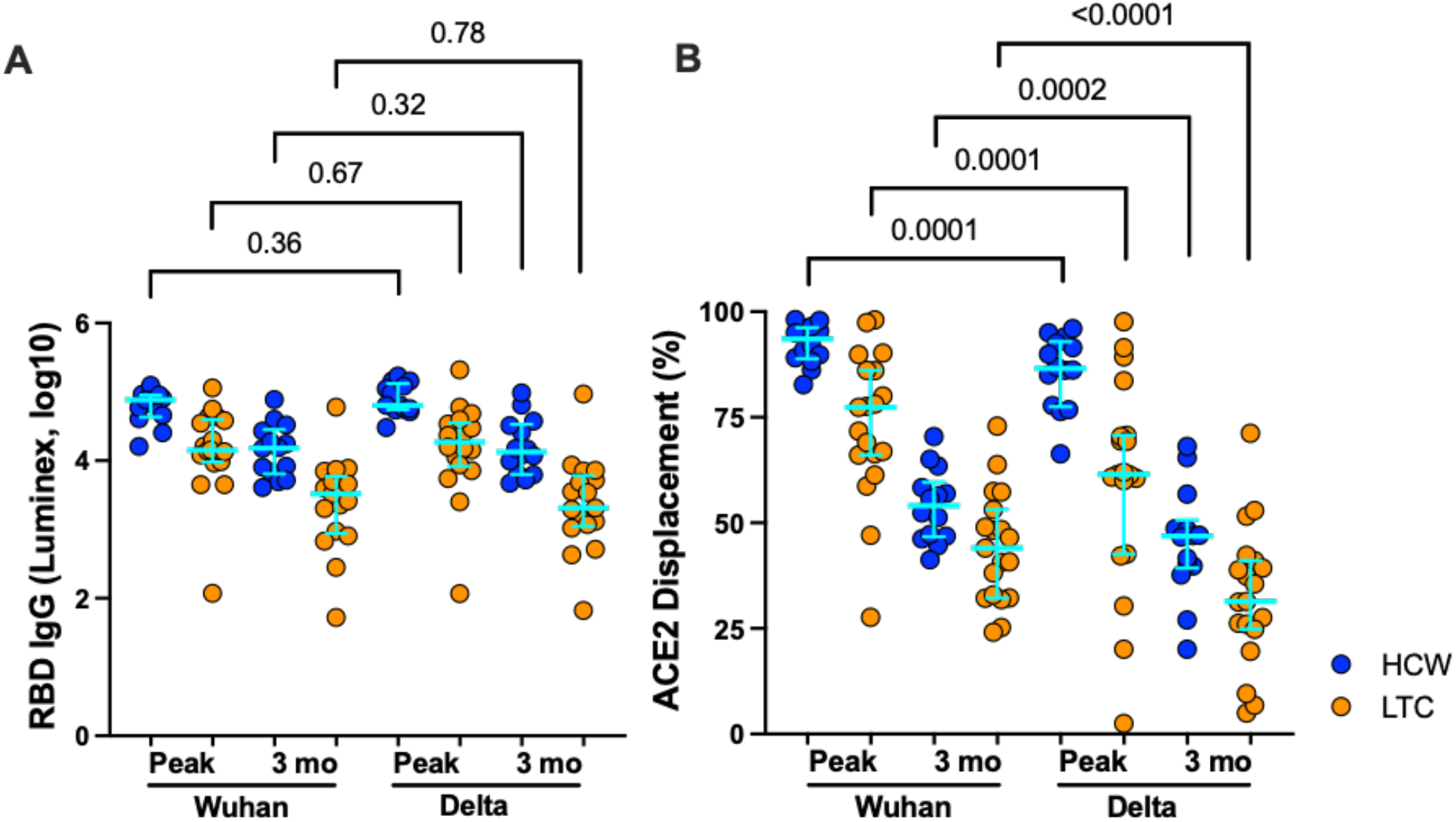
Antibody responses against the Delta variant after two doses of mRNA vaccine. *Panel A:* Binding IgG antibody responses to spike RBD from the original Wuhan strain and the Delta variant, measured by Luminex ELISA, one month following the second vaccine dose (peak) and three months after this dose (3 mo) in a subset of HCW (blue circles) and individuals living in long term care or assisted living facilities (LTC; orange) who were COVID-19 naive at study entry. Bars represent median and IQR, though p-values are computed using the Wilcoxon paired test as the measurements are paired. *Panel B*: ACE2 competition results, measured by Luminex ELISA, in the same individuals.

## DISCUSSION

This study extends our understanding of the magnitude and durability of antibody responses induced by COVID-19 mRNA vaccines across the adult age spectrum ^23-27^. Overall, responses in older adults are impaired both quantitatively (i.e., fewer binding antibodies) and functionally (i.e., lower ACE2 displacement and neutralization activity) compared to younger adults, even after two vaccine doses. Importantly, multivariable analyses confirmed older age as an independent determinant of poorer immune responses following one or both vaccine doses, even after controlling for chronic health conditions that can accumulate with age and compromise immunity ^15-17^. Moreover, though the number of chronic health conditions was also independently associated with lower binding antibody titers and male sex was independently associated with lower ACE2 displacement activity after one dose, the negative impact of these variables on vaccine responses diminished after two doses. Our results thus identify age as the most critical variable modulating antibody responses after COVID-19 mRNA vaccination.

We also explored the durability of the humoral response at three months following the second vaccine dose in a subset of HCW (median age 38 years) and LTC residents (median age 82 years). Despite this timepoint being only two months after we measured peak immune responses, antibodies had waned significantly in both groups: indeed, assuming that decay occurred exponentially, we estimate the half-life of anti-RBD binding antibodies to be 37 days [95% CI 32-42] in our study participants. This suggests that antibody durability following mRNA vaccination is substantially lower compared to that following infection, which was estimated to be ∼116 days in a recent study of convalescent individuals ^28^. Regardless, humoral responses remained overall substantially lower among LTC residents at all timepoints tested, and the “diminished” binding antibody levels observed in HCW at three months following the second vaccine dose were comparable to “peak” levels seen in LTC at one month following the second dose, allowing us to contextualize the extent of immune disadvantage in older adults. Of note, while antibody recognition of the B.1.617.2 (Delta) variant RBD was similar to that of the Wuhan strain, ACE2 competition activity against the Delta RBD was universally reduced, suggesting that older adults will remain more susceptible to infection by this variant at all stages after vaccination due to their weaker overall responses compared to younger individuals.

Our observations are consistent with evidence from other infections describing poorer immune reactivity among older adults that can be mitigated in part by modifying vaccine formulations (e.g., by increasing antigen concentrations or additional adjuvants) or providing booster immunizations more frequently ^15-17^. Recent reports from the UK ^29^ and Germany ^30^ have also demonstrated age-related impairments in binding and neutralizing antibodies following immunization with the COVID-19 mRNA vaccine BNT162b2, though T cell responses were more similar between younger and older participants. However, these studies did not examine the durability of vaccine-induced immune responses in older adults, which is of paramount importance as more people complete the current two-dose vaccine schedule. Indeed, recent increases in SARS-CoV-2 infections among doubly vaccinated individuals ^31^, including outbreaks in LTC facilities ^14^, underscores this ongoing risk.

Our observation that 35% of older adults failed to neutralize SARS-CoV-2 (USA-WA1/2020 strain) even after two vaccine doses also emphasizes the ongoing infection risk in this population. Furthermore, while we did not perform virus neutralization assays using the Delta variant, our ACE2 competition results suggest that neutralization activity against Delta will be even lower than that against the Wuhan strain. Given the ability of SARS-CoV-2 variants to evade at least some aspects of vaccine-elicited immunity ^20^, our results support the prioritization of older adults for receipt of supplemental vaccine doses.

A limitation of our study is that precise immune correlates of protection for SARS-CoV-2 transmission and disease severity remain incompletely characterized ^32^, so the implications of our results as they relate to individual-level control of COVID-19 remain uncertain. The levels of immunity induced in older adults seen here following vaccination may be sufficient to prevent symptomatic infection or severe disease in many cases, so studies linking vaccine immunogenicity data to clinical outcomes specifically among older adults are needed. Due to the timing of the vaccine rollout in British Columbia, our sample size for the durability assessments was modest; however, the weaker responses observed among LTC residents were nevertheless statistically significant. Due to small numbers of participants who received Moderna, we were unable to assess differences in responses between mRNA vaccines ^14,33^.

Overall, our results extend a growing body of evidence indicating that COVID-19 mRNA vaccines are less immunogenic in older compared to younger adults and reveal substantial waning of immunity across all ages in the first three months following receipt of two doses of vaccine. The combined effects of lower peak immunity and natural waning of vaccine-induced responses may leave older adults at continued risk of infection by SARS-CoV-2 or its variants.

## Supporting information

Supplemental Files_combined

## Data Availability

As per funder requirements, data will be deposited into a national database hosted by the Canadian COVID-19 Immunity Task Force (CITF) by the study end date (Spring 2022). Prior to then, data are available to interested researchers upon reasonable request to the corresponding authors.

## ACKNOWLEDGEMENTS

We thank the leadership and staff of Providence Health Care, including long-term care and assisted living residences, for their support of this study. We thank the phlebotomists and laboratory staff at St. Paul’s Hospital, the BC Centre for Excellence in HIV/AIDS and Simon Fraser University for assistance. Above all, we thank the participants, without whom this study would not have been possible.

