## Supplemental Files_combined for "Reduced magnitude and durability of humoral immune responses by COVID-19 mRNA vaccines among older adults"

**Supplemental Table 1. Multivariable Analyses (Roche Elecsys)**

| Immunogenicity outcome | Variable | Time point | | | |
|---|---|---|---|---|---|
| | | 1 month after 1st dose | | 1 month after 2nd dose | |
| | | β estimate (95% CI) | p | β estimate (95% CI) | p |
| Log10 RBD Total Ig | Age | -0.0087 (-0.015 to -0.0024) | **0.007** | -0.0073(-0.013 to -0.0019) | **0.009** |
| | Male Sex | -0.10 (-0.23 to 0.21) | 0.9 | 0.026 (-0.16 to 0.21) | 0.8 |
| | White ethnicity | -0.049 (-0.27 to 0.17) | 0.7 | 0.15 (-0.036 to 0.34) | 0.1 |
| | # Chronic health conditions | -0.14 (-0.23 to -0.043) | **0.005** | -0.12 (-0.21 to -0.042) | **0.003** |
| | Moderna vaccine (vs. Pfizer) | -0.23 (-0.67 to 0.19) | 0.3 | 0.19 (-0.15 to 0.53) | 0.3 |
| | Days after vaccine dose | -0.0022 (-0.041 to 0.037) | 0.9 | 0.015 (-0.020 to 0.050) | 0.4 |

CI, confidence interval

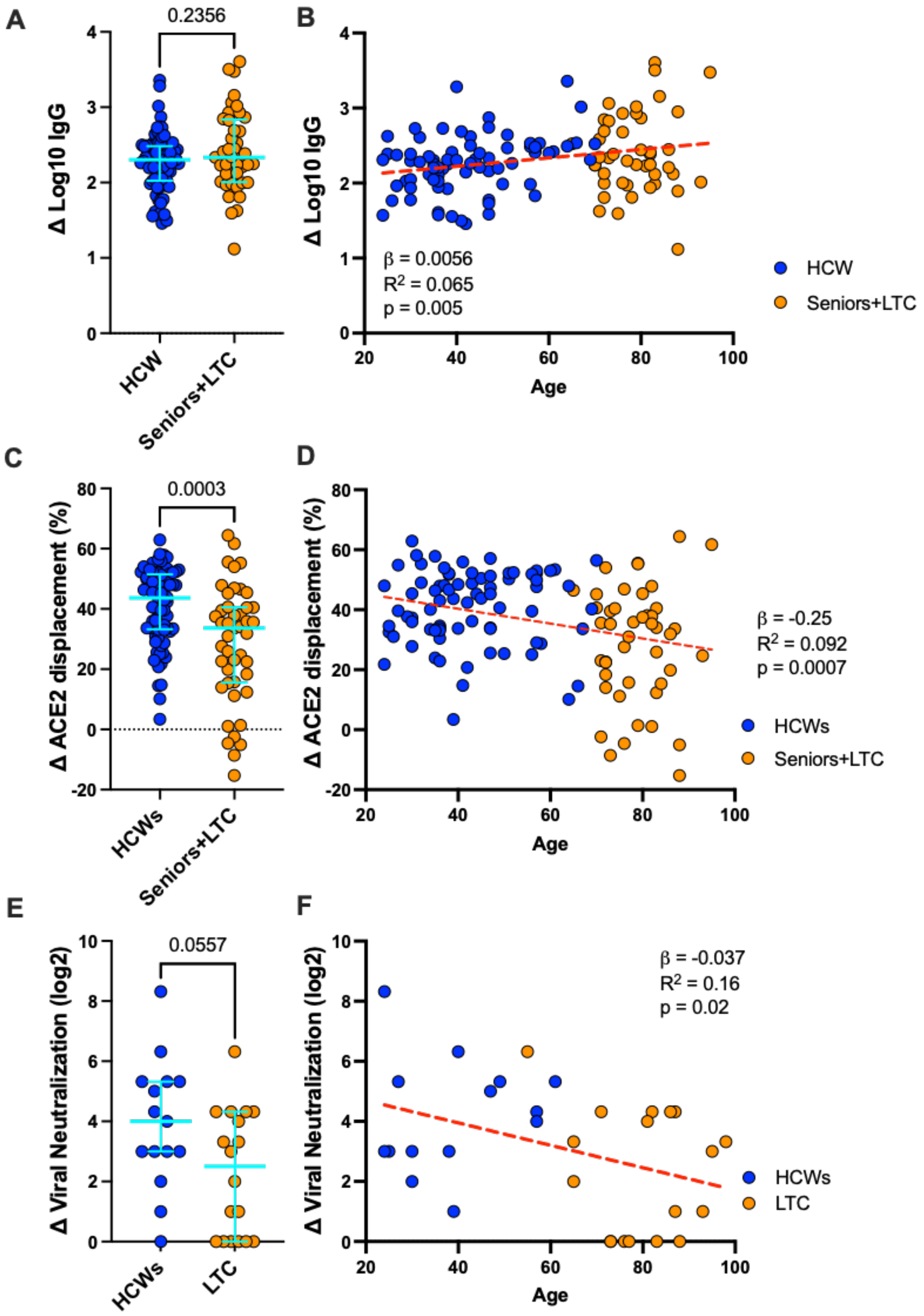


**Supplemental Figure 1: Impact of second vaccine dose on immune responses.**

**Supplemental Figure 1: Impact of second vaccine dose on immune responses.** *Panel A*: Increase in log10 binding IgG antibody responses to RBD in plasma after the second vaccine dose, measured by Luminex ELISA, in HCW (blue circles) and Seniors+LTC (orange circles) who were COVID-19 naive at study entry. Bars represent median and IQR. P-value computed using the Mann-Whitney U-test. *Panel B:* Same data as A, but plotted by age. Statistics and linear model (red line) computed using ordinary least-squares regression. *Panel C*: Increase in ACE2-displacement function of vaccine-induced antibodies after the second dose, in the same participants. *Panel D:* Same data as C, but plotted by age. *Panel E:* Fold-increase in viral neutralization ability of vaccine-induced antibodies after the second dose in a subset of participants who were COVID-19 naive at study entry. *Panel F:* same data as E, but plotted by age.

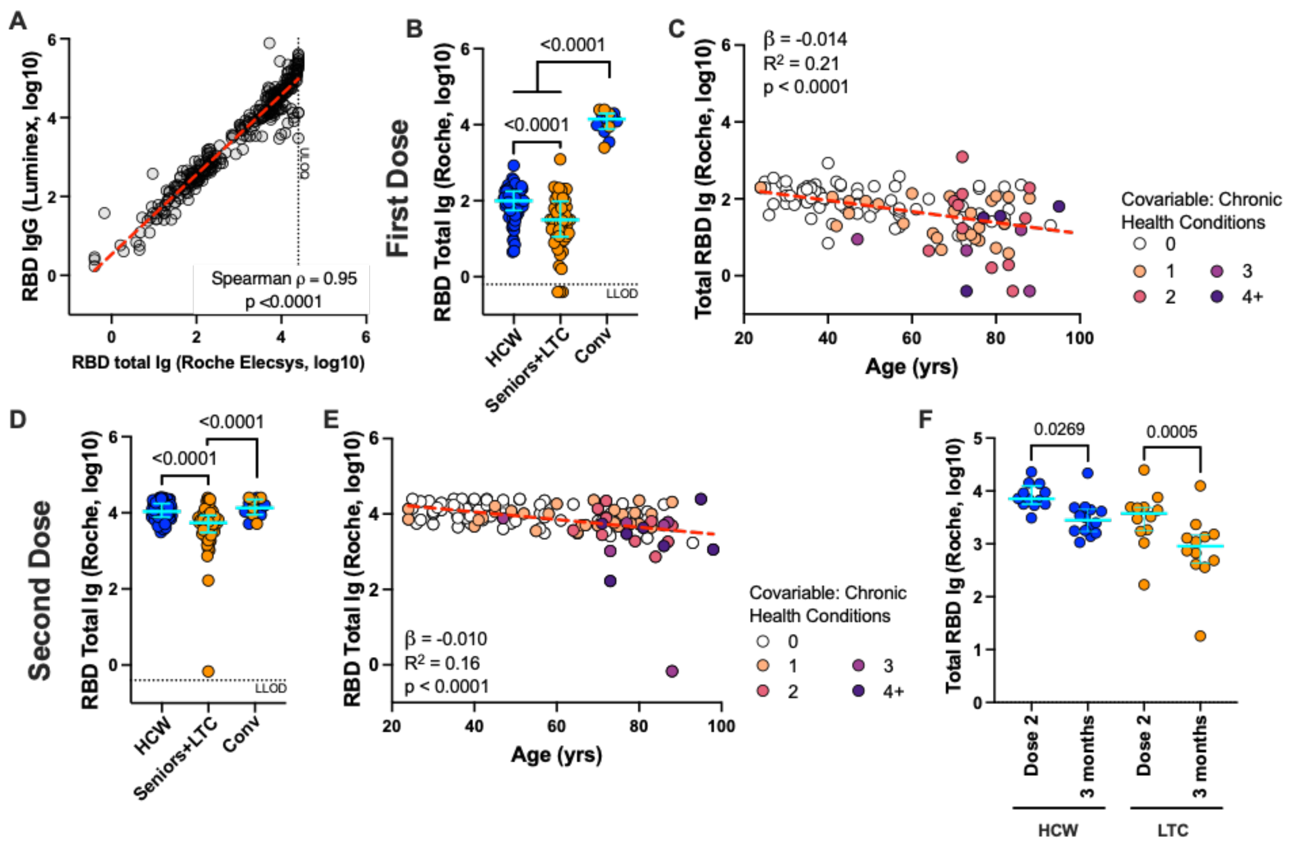


**Supplemental Figure 2: Binding antibody responses to RBD following first and second vaccine doses, measured using a commercial assay.**

**Supplemental Figure 2: Binding antibody responses to RBD following first and second vaccine doses, measured using a commercial assay.** *Panel A*: Spearman's correlation between RBD IgG responses measured in plasma by Luminex ELISA and total antibody responses measured in serum using the Roche Elecsys SARS-CoV-2 S assay. ULOQ = Upper limit of quantification in the Roche assay, which was >25,000 U/mL; 11 samples had measurements above this limit. *Panel B*: Binding antibody responses to the RBD as measured by the Roche assay following one vaccine dose in HCW (blue circles) and Seniors+LTC (orange circles) who were COVID-19 naive at study entry, and COVID-19 convalescent participants (Conv; colored as above). Bars represent median and IQR. P-values computed using the Mann-Whitney U-test. LLOD = Assay lower limit of detection. *Panel C*: Same data as the HCW and Seniors+LTC groups shown in panel B, but plotted by age. Statistics and linear model (red line) computed using ordinary least-squares regression. *Panels D and E:* same as B and C, but after two doses of mRNA vaccine. *Panel E:* Binding antibody responses as measured by the Roche assay, one month following the 2nd vaccine dose (Peak) and three months after this dose (3 months) in a subset of HCW (blue circles) and individuals living in long-term care or assisted living facilities (LTC; orange) who were COVID-19 naive at study entry. P-values computed using the Wilcoxon paired test.
